# A new Far-UVC based method for germ free hospitals and travel: Initus-V

**DOI:** 10.1101/2021.04.23.21255969

**Authors:** Ayhan Olcay, Serdar Baki Albayrak, İbrahim Faruk Aktürk, Mehmet Cengiz Akbülbül, Onur Yolay, Hande İkitimur, Mehmet Çetin Bayer

## Abstract

Direct contact and airborne spread are main mechanisms of transmission for SARS-CoV-2, and virus can stay viable for at least 3 hours in aerosols. Initus-V system uses a Far-ultraviolet C (UVC) system, UVC resistant textile and googles to provide virus, bacteria and spore free environments in hospitals, crowded public places and travel environments. Initus-V system may help in prevention of epidemic diseases such as Coronavirus disease-19 (Covid-19), influenza, treatment of airborne viral diseases and spread of hospital-borne resistant infections.

## Introduction

Coronavirus disease-19 (Covid-19) first appeared in December 2019 was first reported in December 2019 and pronounced as pandemic March 11, 2020. Due to inexperience in global pandemics Covid-19 has spread to 5.3 million confirmed cases and killed over 340,000 patients as of May 25, 2020 (1). Transmission of beta coronavirus causing COVID-19 is through both direct contact and airborne routes (2). Due to up to survival of virus in aerosols up to 3 hours and asymptomatic carriers new technologies are essential to prevent spread of the disease in hospitals and public places (3). Ultraviolet (UV) light exposure is effective against airborne viruses and low pressure mercury-vapor arc lamps and xenon lamps are commonly used as 254nm or broad spectrum UV sources (4-6). These UV lights cannot be used in the presence of living organisms due to serious skin and eye problems (7-10).

Ultraviolet germicidal irradiation (UVGI) is currently used in high risk settings (11-13). UVGI effectively can kill different microorganisms in various settings (13-15). UV can be used for air disinfection either as duct irradiation or upper room UVGI (UV irradiation above people’s heads in a room). UVGI was effectively used in tuberculosis wards in the past but it has recently become popular during Covid-19 pandemic (16,17).

There is a recent development in UVC field called far-UVC. Far-UVC light (207 to 222 nm) is a limited spectrum of UVC and is as efficient as conventional germicidal UV light in killing microorganisms (18) and recent revealed that Far-UVC exposure do not cause skin and eye problems as in conventional UVC (19-22). Far-UVC light can only penetrate a few micrometers in biologic materials and startum corneum and cannot reach living human cells in the skin or eyes, being absorbed in the skin stratum corneum or the ocular tear layer. A few micrometer of biological penetrance is enough for killing virus and bacteria. Far-UVC has same germicidal efficacy but without health hazards of conventional UVC (19-22). Far-UVC light (207 or 222 nm) generated by inexpensive excimer lamps which can be deployed in occupied public locations (23-25). Low-dose-rate far-UVC can be a good tool to prevent spread of aeorosolized viruses in public locations (26).

Initus-V system uses a Far-UVC system, UVC resistant textile and googles to provide virus, bacteria and spore free environments in hospitals, crowded public places and travel environments. Initus-V system may help in prevention of epidemic diseases such as Covid-19, influenza, treatment of airborne viral diseases and spread of hospital-borne resistant infections.

## Methods and Results

### 222nm UVC light source

Initus-V’s 222nm light source was produced by Vestel Inc. (Manisa, Turkey) and InnowayRG Inc. (Istanbul, Turkey) consists of a krypton-chloride (Kr-Cl) excimer lamp and an optical filter with emission maximum output at wavelength 222 nm limiting the emission wavelengths to 200-230 nm. The lamp unit consists of a lamp, air cooling fan, mirrors and a special band-pass filter. The attached filter was used to remove almost all other wavelengths from the spectrum except the dominant 222 nm emission wavelength. The intensity of 222 nm light was measured using an S-172 / UIT250 accumulated UV meter (Ushio Inc.) and was found to be maximum 2.23 uW/cm^2^ at center and minimum 1.34 uW/cm^2^ from 2.5 mt distance (**Figure 1**).

**Figure 1.**
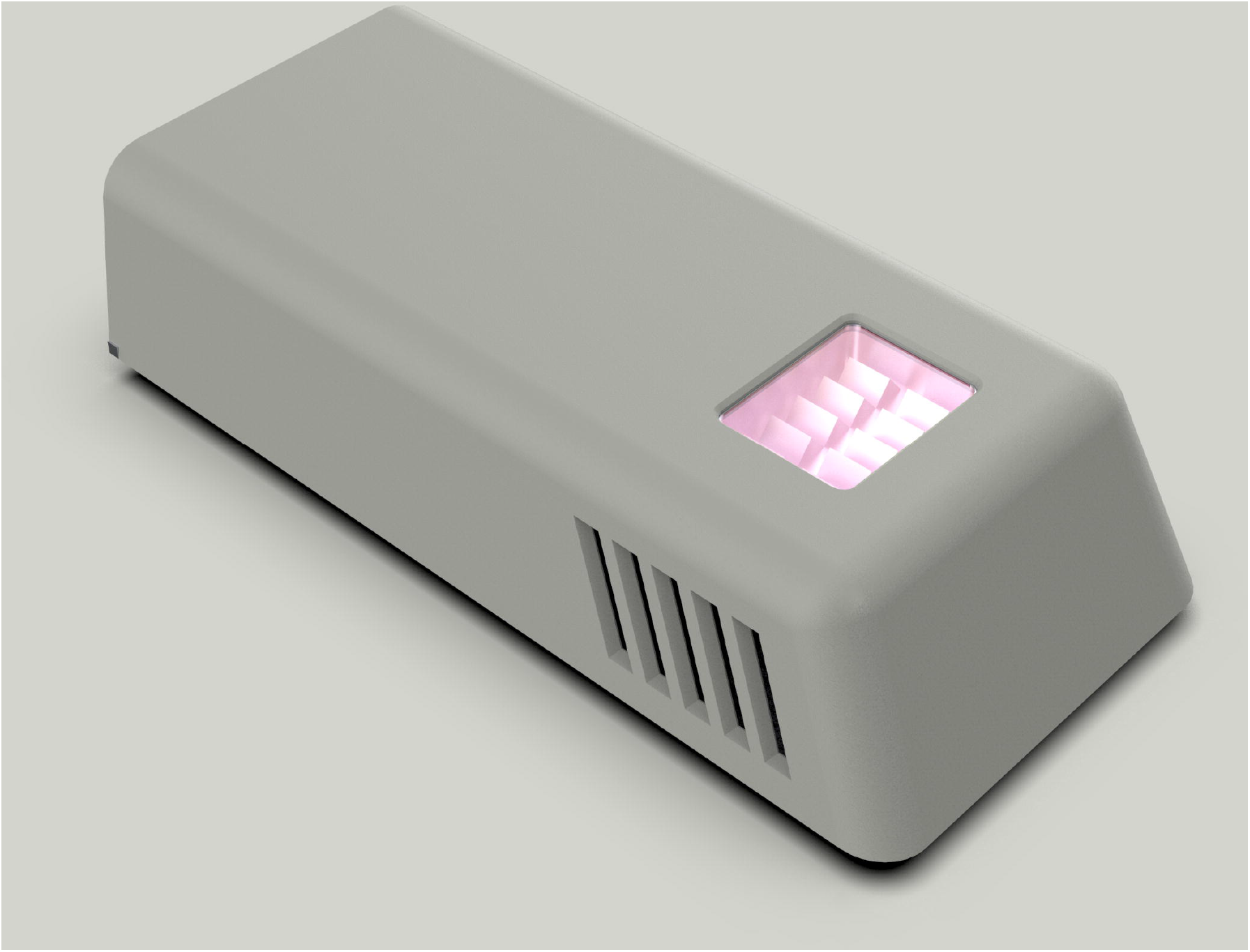
Initus-V’s 222 nm Far-UVC light source produced by Vestel and InnowayRg Inc.

### Daily allowable 222nm UVC radiation limits

The limit for people exposed to 222 nm wavelength UVC in the 8-hour period determined by International Commission on Non-ionizing Radiation Protection) (ICNIRP) is approximately 22 mJ / cm^2^ (27).

### Textile Fabric used in Initus-V system and UVC 222 nm transmittance measurements

We measured by Mettler Toledo UV7 spectrophotometer device 222 nm wavelength transmittance of the textile fabric used in Initus-V overalls and it was 1.5% on avarage. Ultraviolet Protection Factor (UPF) is an indicator of how much UV radiation (both UVB and UVA) a fabric allows to reach your skin. UPF 50 fabric blocks 98 percent of the sun’s rays and allows two (1/50) percent to penetrate, thus significantly reducing your exposure risk Textile fabric used in Initus-V system is in the UPF 50+ class **(Figure 2)**.

**Figure 2.**
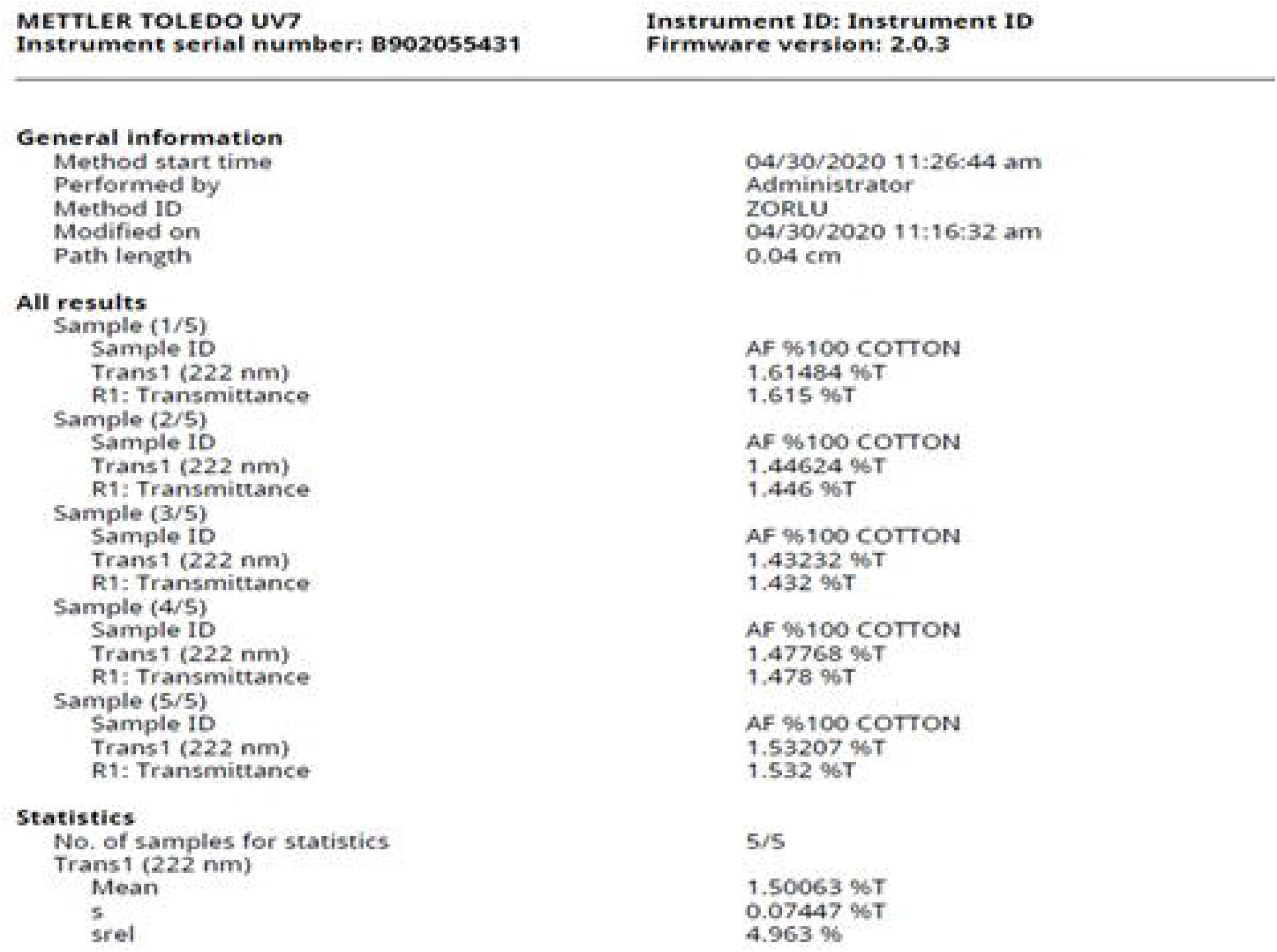
Initus-V textile fabric overall 222nm wavelength transmittance measurements by Mettler Toledo UV7 spectrophotometer

### Installation of Initus-V 222nm UVC source in the working environment and calculations of UVC to be exposed

In order for the UVC exposure of a 170 cm operator under textile fabric overalls to remain within legal limits, Initus-V UVC sources should be placed 220 cm and above from the ground. With a design that will remain within the daily allowable limits of 22 mj/cm^2^ with 1.5% textile fabric permeability to the head area, UVC irradiation can be from top or sides. It corresponds to 22 mj/cm^2^ in 8 hours under the fabric and 1488 mj/cm^2^ on the surface of textile fabric. These values are reached at a distance of 50 cm from Initus-V UVC source and devices must be mounted >50 cm from nearest body part of the operator.

### Operating Room/Intensive Care System setup where personnel can work inside with an active Initus-V Far-UVC light

In Initus-V system, 200-222nm UVC light source is used together with UVC-proof glasses and textile fabrics while UVC is active in the environment. It is possible to get rid of viruses, microbes, bacteria and pathogens that threaten human health by using the Initus-V system. Despite many papers documenting safety of Far-UVC light on skin and eyes we preferred textile and google protection to stay within regulatory limits of ICNIRP. There no long term human studies with far-UVC technology and regulatory bodies are very hesitant to allow use of far-UVC in the presence of living subjects in environment. Patent applications were submitted to Turkish Patent Institute for Initus-V system and our solution provides immediate use of Far-UVC technology without any regulatory body conflict or restriction. Textile innovation keeps human exposure to 222 nm UVC within regulatory limits and makes far-UVC usable in all environments without waiting for long term human studies.

### Initus-V System Description with Figures

The operating room, intensive care, emergency room or environments requiring sterility are illuminated with fixed Initus-V UVC light sources that emit light in the wavelength range of 200-222 nm or movable UVC lamps to prevent surgical field infection (**Figure 3**). Operators and other auxiliary personnel wear UVC-protective overalls and UVC-protected glasses during the procedure. UVC-protecting overalls are in the form covering whole body and head and zippered in the front or as poncho for easier movement and comfort. If the patient is in the intensive care unit or operating room, it is covered with a UVC protective cover (**Figure 4**,**5**). The operator can perform the operation while the UVC lamps are working actively and any virus or bacteria in the environment is inactivated by UVC.

**Figure 3.**
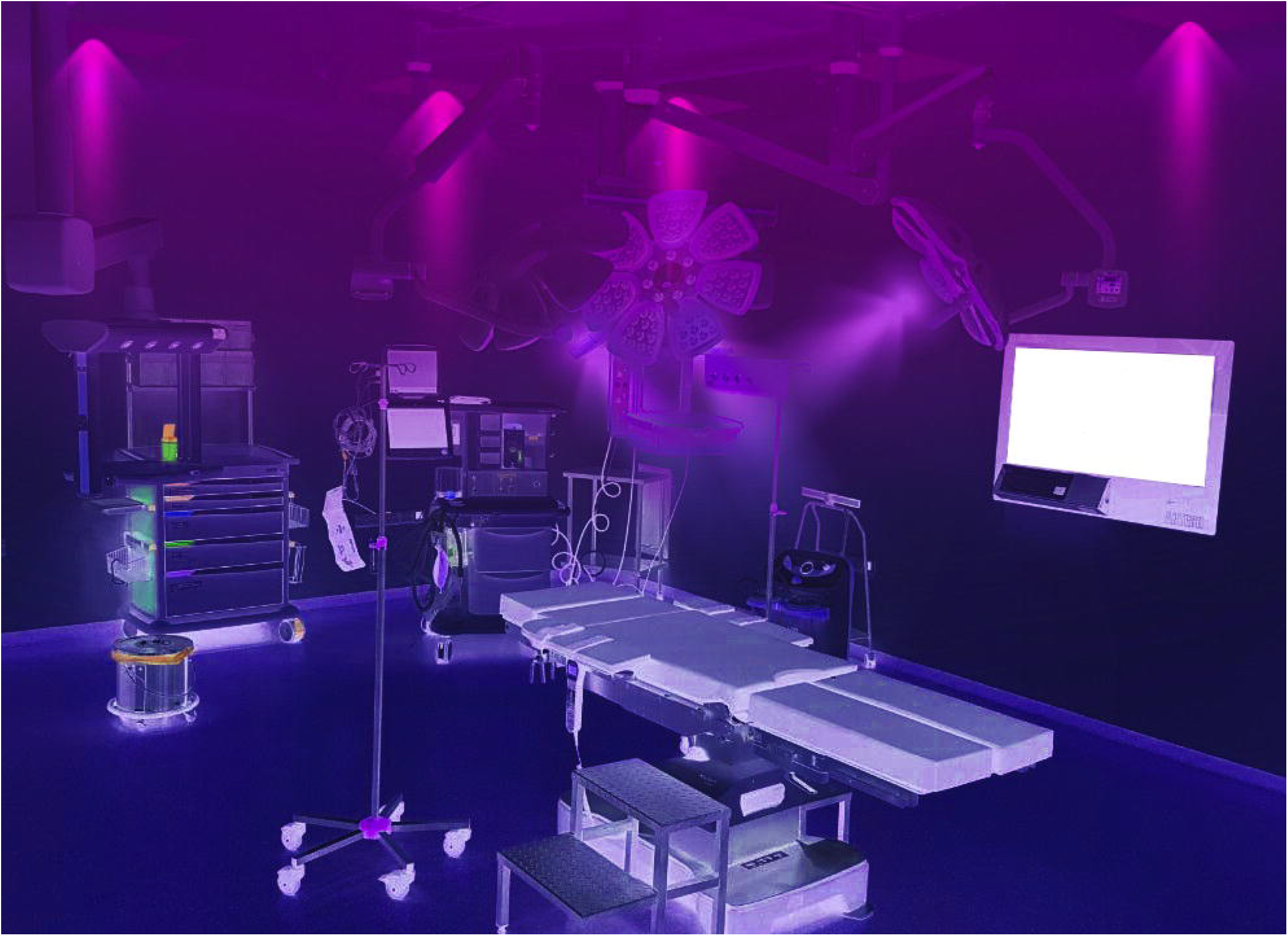
The operating room requiring sterility are illuminated with fixed Initus-V UVC light sources that emit light in the wavelength range of 200-222 nm or movable UVC lamps to prevent surgical field infection

**Figure 4.**
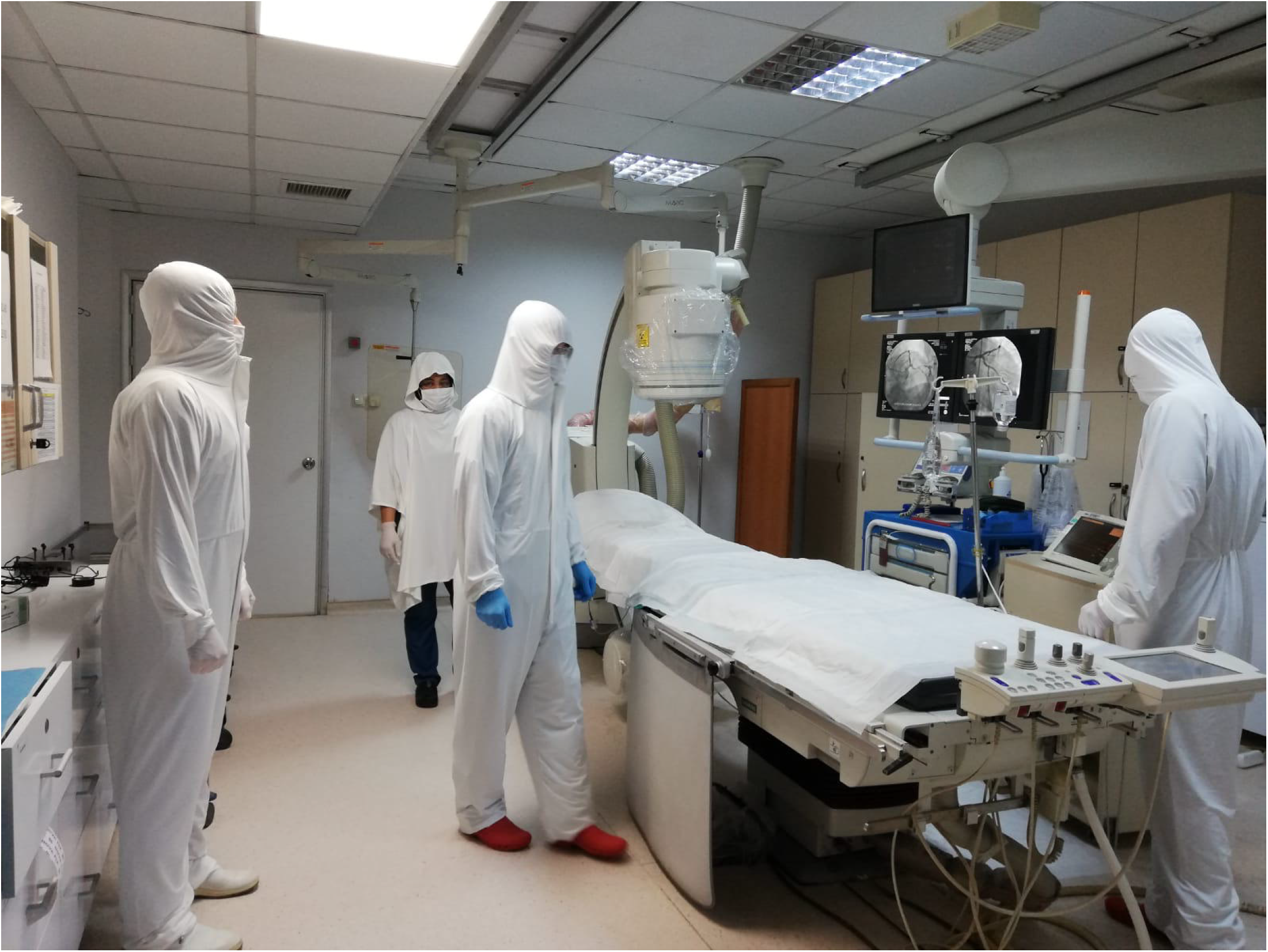
Operators and other auxiliary personnel wear UVC-protective overalls in the form covering whole body and zippered in the front or as poncho for easier movement and comfort.

**Figure 5.**
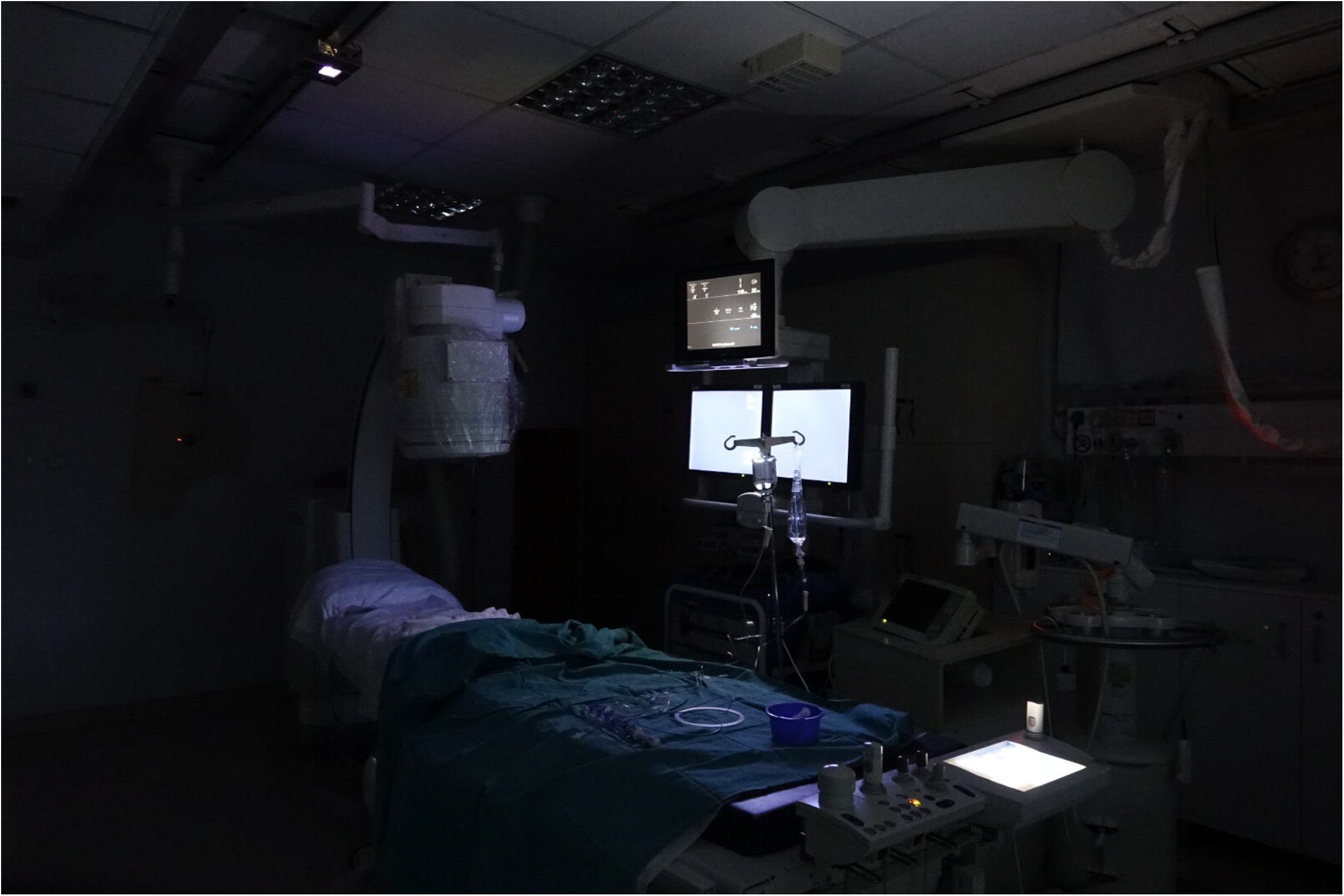
An angiography suit illuminated with Initus-V UVC light souce for exposure measurements before patient clinical study.

## Discussion

Infection protection equipments such as protective overalls, glasses, gloves, masks against viruses and bacteria only prevent human contact without eliminating viruses and bacteria in the air. Initus-V system may reduce the use of protective equipment and provide a more comfortable working environment for employees. Factories, airports, shopping malls, schools, buses, airplanes and similar areas that cannot be used due to social distance or infection threat may be able to reduce social distance and normalize production, working and travel conditions with Initus-V system use. Far-UVC 200 nm light transmission from the cornea to the lens is predicted to be essentially zero. It has been observed that 207 nm UV light kills MRSA efficiently, but unlike conventional germicidal UV lamps, it kills very few cells in human cells (19-22). There was no tumor induction in Xpa□knockout mice and wild□type mice by repetitive irradiation with 222□nm UVC, using a tumor producing protocol which was shown to produce tumor when irradiated with broad□band UVB (23)

In a three-dimensional human skin model, 207-nm UV light produced virtually no mutagenic UV-associated DNA lesions, unlike significant side effects caused by a traditional germicidal UV lamp (24,25). In this way, UVC sterilization is aimed without harming human health when there are people in the environment.

In a previous study effect of exposure of the skin to UV radiation of wavelength 222 nm using a device that is used to sterilize equipment (Sterilray™ Health Environment Innovations, Dover, New Hampshire, USA) was assessed by induction of skin erythema and induction of cyclopyrimidine dimers (CPD) in skin biopsies. Study found that not only is erythema inducible using the UVC emitting Sterilray, but also that DNA damage in the form of the skin cancer– associated CPDs, has also been induced. These events are occurring at Sterilray dosage levels below the threshold for bacteriostatic/bacteriocidal effects suggesting that frequent, several times daily, use of Sterilray irradiation is unlikely to be tolerated as a non-chemical antiseptic for human skin (28).

A recent paper revealed that viable SARS-CoV-2 can be present in aerosols generated by a Covid-19 patient in a hospital room in the absence of an aerosol-generating procedure, and can thus serve as a source for transmission of the virus in this setting (29). Air samples were collected in a room that was part of a designated Covid-19 ward. The room had six air changes per hour and the exhaust air underwent triple filter treatment, coil condensation (to remove moisture), and UV-C irradiation prior to recycling 90% of the treated air back to the room. The air-samplers were stationed from 2 to 4.8 m away from the patients. This article suggests that despite maximum air filtering aerosol transmission is possible from a 4.8 mt distance of a patient and Initus-V system can inactivate airborne viruses regardless of ventilation.

Low-dose-rate far-UVC can be a good tool to prevent spread of aeorosolized viruses in public locations but regulations about UVC exposures are very strict and using Far-UVC with UVC resistan textile like in Initus-V seems to be only way to introduce this technology into common public places without waiting for long term human trial results or regulatory changes (26)

Contrary to the current technique, UVC systems with a wavelength of 200-222 nm are low in error, do not harm human health, are easy to install, effective and not dependent on consumables. By combining this system with clothes and covers made of UVC-proof fabric in Initus-V, we eliminate the long-term damage hesitation and enable working under UVC light immediately without any concern. Our recommended system includes UVC-protected wearable fabric and clothing, patient and operating room cover, UVC-proof glasses, and laboratory lighting in the 200-222 nm range.

## Data Availability

All referred data is available

## References

1. World Health Organization. Coronavirus disease (COVID-2019) situation reports. Available on: https://www.who.int/emergencies/diseases/novel-coronavirus-2019/situation-reports (2020).

2. van Doremalen, N. et al.. Aerosol and surface stability of sars-cov-2 as compared with sars-cov-1. N. Engl. J. Med, (2020).

3. Bai, Y. et al.. Presumed asymptomatic carrier transmission of covid-19. JAMA, (2020).

4. Kowalski, W. J. Ultraviolet Germicidal Irradiation Handbook: UVGI for Air and Surface Disinfection. New York: Springer, (2009).

5. Budowsky, E. I. et al.. Principles of selective inactivation of viral genome. I. UV-induced inactivation of influenza virus. Arch. Virol. 68(3-4), 239–47 (1981).

6. Naunovic, Z., Lim, S. & Blatchley, E. R. III. Investigation of microbial inactivation efficiency of a UV disinfection system employing an excimer lamp. Water Res. 42(19), 4838–46 (2008).

7. Trevisan, A. et al.. Unusual high exposure to ultraviolet-C radiation. Photochem. Photobiol. 82(4), 1077–9 (2006).

8. Zaffina, S. et al.. Accidental exposure to UV radiation produced by germicidal lamp: case report and risk assessment. Photochem. Photobiol. 88(4), 1001–4 (2012).

9. Setlow, R. B. et al.. Wavelengths effective in induction of malignant melanoma. Proc. Natl Acad. Sci. USA 90(14), 6666–70 (1993).

10. Balasubramanian, D. Ultraviolet radiation and cataract. J. Ocul. Pharmacol. Ther. 16(3), 285–97 (2000).

11. Green, C. F.; Scarpino, P. V.; Jensen, P.; Jensen, N. J.; Gibbs, S. G., Disinfection of selected Aspergillus spp. using ultraviolet germicidal irradiation. Can. J. Microbiol. 2004, 50 (3), 221–4.

12. Xu, P.; Kujundzic, E.; Peccia, J.; Schafer, M. P.; Moss, G.; Hernandez, M.; Miller, S. L. Impact of environmental factors on efficacy of upper-room air ultraviolet germicidal irradiation for inactivating airborne mycobacteria. Environ. Sci. Technol. 2005, 39 (24), 9656–64.

13. Xu, P.; Peccia, J.; Fabian, P.; Martyny, J. W.; Fennelly, K.; Hernandez, M.; Miller, S. L. Efficacy of ultraviolet germicidal irradiation of upper-room air in inactivating bacterial spores and mycobacteria in full-scale studies. Atmos. Environ. 2003, 37, 405–19.

14. Ko, G.; First, M. W.; Burge, H. A. The characterization of upper room ultraviolet germicidal irradiation in inactivating airborne microorganisms. Environ. Health Perspect. 2001, 110 (1), pp. 95–101.

15. Perkins, J. E.; Bahlke, A. M.; Silverman, H. F. Effect of ultraviolet irradiation of classrooms on spread of measles in large rural central schools. Am. J. Public Health Nations Health 1947, 37, 529–37.

16. Dumyahn, T.; First, M. Characterization of ultraviolet room air disinfection devices. Am. Ind. Hyg. Assoc. J. 1999, 60, 219–27.

17. CDC, Guidelines for preventing the transmission of Mycobacterium tuberculosis in health-care facility. MMWR 1994, 43 (RR-13), 1–132

18. Narita, K. et al.. 222-nm UVC inactivates a wide spectrum of microbial pathogens. J Hosp Infect (2020).

19. Buonanno, M. et al.. 207-nm UV light - a promising tool for safe low-cost reduction of surgical site infections. I: in vitro studies. Plos One 8(10), e76968 (2013).

20. Buonanno, M. et al.. 207-nm UV light-a promising tool for safe low-cost reduction of surgical site infections. II: In-Vivo Safety Studies. PLoS One 11(6), e0138418 (2016).

21. Ponnaiya, B. et al.. Far-UVC light prevents MRSA infection of superficial wounds in vivo. Plos One 13(2), e0192053 (2018).

22. Narita, K. et al.. Disinfection and healing effects of 222-nm UVC light on methicillin-resistant Staphylococcus aureus infection in mouse wounds. J. Photochem. Photobiol. B 178(Supplement C), 10–18 (2018).

23. Yamano, N. et al.. Long-term effects of 222 nm ultraviolet radiation C sterilizing lamps on mice susceptible to ultraviolet radiation. Photochem Photobiol, (2020).

24. Narita, K. et al.. Chronic irradiation with 222-nm UVC light induces neither DNA damage nor epidermal lesions in mouse skin, even at high doses. PLoS One 13(7), e0201259 (2018).

25. Buonanno, M. et al.. Germicidal efficacy and mammalian skin safety of 222-nm uv light. Radiat. Res. 187(4), 483–491 (2017).

26. Buonanno M, Welch D, Shuryak I, Brenner DJ. Far-UVC light (222□nm) efficiently and safely inactivates airborne human coronaviruses. Sci Rep. 2020;10(1):10285. Published 2020 Jun 24. doi:10.1038/s41598-020-67211-2

27. https://www.icnirp.org/cms/upload/publications/ICNIRPUV2004.pdf

28. Woods JA, Evans A, Forbes PD, et al. The effect of 222-nm UVC phototesting on healthy volunteer skin: a pilot study. Photodermatol Photoimmunol Photomed. 2015;31(3):159–166. doi:10.1111/phpp.12156

29. John A Lednicky, Michael Lauzardo, Z. Hugh Fan, Antarpreet S Jutla, Trevor B Tilly, Mayank Gangwar, Moiz Usmani, Sripriya N Shankar, Karim Mohamed, Arantza Eiguren-Fernandez, Caroline J Stephenson, Md. Mahbubul Alam, Maha A Elbadry, Julia C Loeb, Kuttichantran Subramaniam, Thomas B Waltzek, Kartikeya Cherabuddi, John Glenn Morris Jr., Chang-Yu Wu. Viable SARS-CoV-2 in the air of a hospital room with COVID-19 patients. medRxiv 2020.08.03.20167395; doi:https://doi.org/10.1101/2020.08.03.20167395

